# Design and evaluation of a streamlined enrichment method for faster co-detection of enteric bacterial pathogens in natural soils and environmental samples

**DOI:** 10.64898/2026.09.23.26363803

**Authors:** Alexis Kapanka, Sharon Kwamutakha, Ellie M. Madson, Collins Ouma, Megan Sinik, Bonphace Okoth, John Agira, Christine S Amondi, Sheillah Simiyu, Fanta D. Gutema, Daniel Sewell, Kelly K Baker

## Abstract

This study developed a streamlined, sensitive and rapid method for co-detection of low concentrations of Salmonella enterica, Shigella flexneri, Listeria monocytogenes, and pathogenic Escherichia coli from soil. First, autoclaved soil from Iowa with serial dilutions of bacteria to confirm individual species recovery rates on selective agar by qPCR under varying primary enrichment times was used. All pathogens were detected by qPCR of colonies recovered from replicate sterile soils spiked with 10^1 or greater bacteria within 5 hours of enrichment. Shiga toxin expressing E. coli (STEC) and L. monocytogenes were consistently co-recovered in autoclaved and natural soil, pathogen spiking concentration, and secondary enrichment time. S.enterica was consistently recovered at autoclaved and natural concentrations, while S. flexneri detection rates fluctuated. Twenty-three soil samples from Kenyan neighborhoods, with prior evidence of contamination, was compared to qPCR detection at each protocol stage: processing, primary enrichment, secondary enrichment, and selective colony isolation. Detection rates substantially increased for S. enterica, STEC and other pathogenic E. coli after enrichment, with optimal detection rates occurring after secondary enrichment. Testing colonies from selective agar resulted in high false negative rates, possibly reflecting competition from native soil microbiota that was absent in spiked sterile soil experiment. Evaluation of the method with other environmental sample types yielded consistent detection patterns. Evidence suggests a protocol including 24-hour primary enrichment, followed by 24-hour secondary enrichment, and qPCR testing of soil DNA at primary and secondary steps is a reproducible and more expedient method for co-detecting common Enterobacteriaceae and L. monocytogenes pathogens in soil.

## INTRODUCTION

In 2021, diarrheal disease was ranked as the fifth leading cause of death and the second leading cause of disability adjusted life years (DALYs) amongst young children, causing around 340,000 (244,000 – 480,000) deaths in children.^1^ Deaths due to diarrhea primarily occur within the Sub-Saharan African and South Asian regions. A wide range of bacterial, viral, and protozoan enteric pathogens cause diarrheal infections.^2,3^ Of bacterial causes, *Shigella spp*., *Salmonella enterica, Campylobacter jejuni*, and pathogenic *Escherichia coli* remain leading bacterial causes of enteric infections that cause moderate to severe diarrhea and diarrhea-related hospitalization and death within heavily affected regions.^4^ Repeated exposure to enteric pathogens can result in adverse long-term outcomes such as environmental enteropathy (EE), wherein increased inflammation and/or microbial pathogenesis cause abnormal intestinal morphology (IM) and impaired barrier function, in addition to defects in linear growth.^5^ This altered IM and function weakens the innate immunity of the gut mucosa against non-commensal enteric pathogens, and thus leaves the host susceptible to re-infection. Identification of pathogen transmission pathways within communities and subsequent implementation of preventative practices can prevent both acute illness and chronic sequalae of infection.

The pathways for transmission of pathogens in human or animal feces through the environment to a new host include, but are not limited to, contaminated food, flies, fingers, soils and surfaces, and fluids.^6^ Soil in domestic and community settings where children play is an important fomite due to its likelihood of fecal contamination from open defecation and feces disposal and its mechanistic role in contamination of other fomites like household food and water supplies, hands and objects.^7^ Young children’s propensity to place objects and hands in their mouth during play and to directly consume soil (geophagy) ^8-10^ has been associated with increased risk of diarrhea and enteropathy.^5,8,11^ Studies in Kenya and Haiti have confirmed high rates of detection for one or more enteric pathogens in soil samples at child play areas,^12-14^ and estimated high exposure doses of one or more enteric pathogens from repeat soil ingestion.^15,16^ These exposures could significantly contribute to the high population prevalence of enteric infections and within-child taxonomic diversity of enteric pathogens detected in humans.^17-19^ Depending on the specific pathogen, soil characteristics, and climate factors, bacteria can persist within soil for months to years, making it an important reservoir in the transmission pathway.^20,21^

Given the evidence on the prevalence of enteric pathogens in soil in diarrheal endemic communities and soil’s potential for causing infections in young children, monitoring soil and fomites containing soil (e.g. surface water) should be an important step in health risk assessments and evaluation of environmental cleanup interventions. There are currently no rapid and sensitive standardized protocols for simultaneous detection and quantification of clinically important enteric pathogens from soil. Many public health studies have evaluated on-site soil contamination using *E. coli* bacterial indicator assays adopted from drinking water safety assessment guidelines, instead of direct quantification of the specific pathogen community. However, these assays can isolate environmental *E. coli*, alongside commensal and pathogenic types, and poorly predict enteric pathogens, making them unreliable indicators of fecal contamination or health risks.^12,14,22-24^ Several culture-dependent, biochemical, and molecular detection methods have been developed for understanding the ecology of soil microbial communities and their roles in soil quality in agricultural settings.^25,26^ However, existing culture-based protocols for detection of enteric bacterial pathogens from soil vary between studies. Some studies detected and enumerated enteric pathogens in soils from agricultural production sites and residential areas using protocols developed for detection of the pathogens from foods.^27-29^ These protocols are labor and/or time intensive, requiring a detection process of more than 72 hours. There is little information regarding the performance of universal primary enrichment and secondary selective enrichment of culture media for detection of enteric bacterial pathogens from soil. Pathogen isolation and confirmation may be complicated by a complex interplay between natural soil phages, bacteria, molds, and fungi and pathogens during the culture process. Other methods used for detection and quantification of enteric bacteria in soil is the direct use of molecular techniques such as PCR and sequencing.^12,14,26,30^ However, detection through this method does not confirm the viability or infectivity of the pathogen, and without a pre-enrichment it can lead to underestimation of the presence and concentrations of the pathogens.^12^

New methods for detecting enteric pathogens in soil are needed to establish the epidemiological linkage of contaminated soil with enteric disease in children. This study aimed to develop and test a streamlined and more rapid method for detection of *Salmonella enterica, Shigella spp*., *L. monocytogenes* and Shiga toxin-producing *E. coli* individually and jointly in soil. Simultaneous detection of multiple enteric pathogens also provides information for better understanding of the diversity of enteric pathogens within the environment, and their potential association with pediatric diarrheal diseases.

## METHODS

### Protocol Development and Experimental design

An experimental study was conducted to evaluate culture-based simultaneous recovery of multiple foodborne pathogens in soil by spiking soil samples with known bacterial strains under different pre-enrichment and selective enrichment incubation conditions. In this experimental study, established protocols for detecting *Salmonella enterica*, pathogenic *E. coli, Shigella* spp., and *L. monocytogenes* from food samples were adapted for bacterial detection in soil samples.^31-34^ The recovery of these pathogens from food samples involves a series of time and labor-intensive procedures that include pre-enrichment with primary enrichment [e.g., Buffered Peptone Water (BPW)] to revive sublethal bacteria, secondary selective enrichment, and selective plating for growing specific bacteria by suppressing other competing background microorganisms. These steps are often followed by confirmatory assays such as biochemical tests and molecular techniques to determine the type of pathogens. We sought to streamline processing steps for co-recovering bacteria using validated enrichment and culture media protocols that have been used for recovery and growth of multiple target bacteria species, for example the use of BPW for recovery of *Enterobacteriaceae* as well as *Campylobacter* spp. from food. While food is often cooked and presumed to lack microbial contamination, soil in real-world conditions contains a complex microbial community of viruses, bacteria, algae, and other life forms that may interfere with pathogen detection for methodological reasons, such as through competition for nutrients and growth on media, or for biological reasons, such as bacteriophagy or lysis. Therefore, our experimental design also evaluated how well-adapted bacteria detection protocols would work in those non-sterile, real-world conditions.

Before evaluating the co-recovery and detection of multiple pathogens, we first performed a set of experiments to identify the culture conditions which maximized recovery of low concentrations of individual pathogen species inoculated into sterile soil while reducing processing steps and time to pathogen detection. Then, we evaluated the simultaneous recovery of these pathogens from three different amounts of sterile (autoclaved) and non-sterile soil samples (0.25, 2.5, and 25 grams (g)) spiked with three different concentrations of known strains of the pathogens. We tested modified protocols of the conventional recovery methods for these pathogens that could shorten protocols, specifically:

1. primary enrichment incubation for 3 hr and 5 hr without a selective enrichment step
2. primary enrichment for 15 hr with selective secondary enrichment for 24 hr
3. primary enrichment for 24 hr with selective enrichment steps for 5 hr and 10 hr

### Soil Sample Collection for Controlled Experiments

For each trial, approximately 200 grams of soil samples were collected from the University of Iowa College of Public Health Multi-Tenant Facility Laboratory campus in Coralville, Iowa. Before use, half of the collected samples were sterilized in an autoclave at 121°C for 15 minutes. In the single pathogen recovery experiment, only the sterile soil samples were spiked and processed while sterile and non-sterile soil were used for multi-pathogen recovery experiments.

### Bacterial Strains and Inoculum Preparation

We used bacterial strains obtained through Biodefense and Emerging Infections Resources (BEI), NIAID, NIH: *Salmonella enterica* subsp. Enterica (Strain ST2380, Serovar Typhimurium, NR-4392), Shiga toxin expressing *Escherichia coli* (STEC) (Strain B2F1, NR-96), *Shigella flexneri* (Strain 2457T, NR-518), and *Listeria monocytogenes* (Strain FSL J1-194, NR-13229) to spike the soil samples. The pathogens were stored in 80% glycerol (Sigma Aldrich, USA) at -80°C until used. One day prior to experimental trials, Salmonella, Shigella, and *E. coli*, were subcultured into 25 ml of Luria-Bertani (LB) broth (MP Biomedicals, France), while *L. monocytogenes* was subcultured into 25 ml of Fraser Broth (OXOID, UK). All broth cultures were placed in a shaking incubator at 37°C for 24 hr. The following day, all the broth cultures were diluted using BPW (OXOID, UK) until an OD600 reading of about 1.0 or 10^8^ colony forming unit (CFU)/ml was obtained using a spectrophotometer. A 10-fold serial dilution of the 108 CFU/ml culture was created in BPW to achieve the final concentrations of 10^4^ CFU/ml, 10^3^ CFU/ml, 10^2^ CFU/ml and 10^1^ CFU/ml. A portion of dilutions were plated on agar to confirm actual concentration in CFU/ml before spiking the soil samples.

### Primary Enrichment Conditions for Pathogen Recovery

In this experiment, four trials were performed to confirm whether we could detect each type of pathogen species from sterile soil samples in monoculture by incubating the aliquots in primary enrichments for 3 and 5 hrs. The 5 hr incubation period was selected based on our previous experiment that demonstrated complete recovery and detection of *Salmonella spp*. from food at a 10^1^ CFU/mL starting concentration.^35^ Briefly, 2.5 g of sterile soil samples were added separately into two sets of five sterile Whirl-Pak bags containing 250 mL of BPW. Each bag was spiked with 100 µl of liquid culture of 10^4^, 10^3^, 10^2^ and 10^1^ CFU/mL resulting in concentrations of 10^3^, 10^2^, 10^1^ and 10^0^ CFU/testing bag. Additionally, 2.5 g of sterile soil was added to another Whirl-Pak bag containing 250 mL BPW to be used as a negative control without bacteria inoculation. All the bags were sealed, homogenized for 2 minutes and incubated at 37°C for 3 and 5 hrs.

To evaluate the effectiveness of the primary enrichment step of bacteria recovery, 100 µl of liquid culture of the primary enrichment from each bag was spread on two selective agar plates for each pathogen targeted in this trial. The selective agar used was Xylose Lysine Deoxycholate (XLD, Remel, USA), MacConkey Agar (BD Difco, France), and Listeria Identification Agar Base (PALCALM, HiMedia, India) for selective growth of *S. enterica* and *S. flexneri*, STEC, and *L. monocytogenes*, respectively. A 100 µl aliquot of the negative control was plated on all selective media. For a positive control, 100 µl of the original liquid bacterial culture at 10^8^ CFU/mL was spread onto a selective agar plate. All plates were placed in an incubator at 37°C for 24 hr. The plates were examined for the presence and absence of growth by observing and characterizing colony morphologies according to manufacturer’s recommendations and comparing them with positive and negative controls.

### Optimization of Primary and Secondary Enrichment for Multi-pathogen Recovery

We designed a new experimental trial based on the results of the single-pathogen recovery experiment to evaluate and compare the detection of multiple pathogens in sterile and non-sterile environmental soil samples of three different amounts by including a selective enrichment step. We evaluated the co-detection of multiple bacteria by increasing the incubation time of the primary enrichment to 15 hr (to optimize *S. flexneri* recovery) followed by selective enrichment for 5 hr and 10 hr periods (Figure 1). Two sets of nine sterile Whirl-Pak bags, one set for sterile soil and the other for non-sterile soil samples were prepared prior to spiking. Within each soil type, three bags were filled with 0.25 g, 2.5 g, and 25 g soil, resulting in three bags per volume per soil type. A 100 µl volume of each pathogen from three different concentrations (10^3^ CFU/ml, 10^2^ CFU/ml, and 10^1^ CFU/ml) was added to each bag resulting in spiking concentrations of 10^2^ CFU, 10^1^ CFU, and 10^0^ CFU per bag, respectively and thus testing concentrations ranging from 10-1/g (10^0^ CFU+25 g soil) to 10^3^/g (10^2^ CFU+0.25 g soil). In this experiment, the concentration of 10^4^ CFU/mL used in the single pathogen recovery test, in which all pathogens were detected in 2.5 g sterile soil samples, was dropped. All the bags were hand agitated for one minute to allow homogenization of the pathogens and soil samples. After homogenization, 250 mL of BPW enrichment broth was added to bags containing 0.25 g and 2.5 g soil, and 225 mL BPW was added to bags containing 25 g soil. For each of the three groups of sample volume, two negative controls (one for sterile and one non-sterile soil samples) were prepared by measuring 2.5 g soil into 250 ml BPW without spiking with any of the pathogens. All the bags containing the pre-enriched samples were sealed, homogenized for 2 minutes and incubated at 37°C for 15 hr.

**Figure 1.**
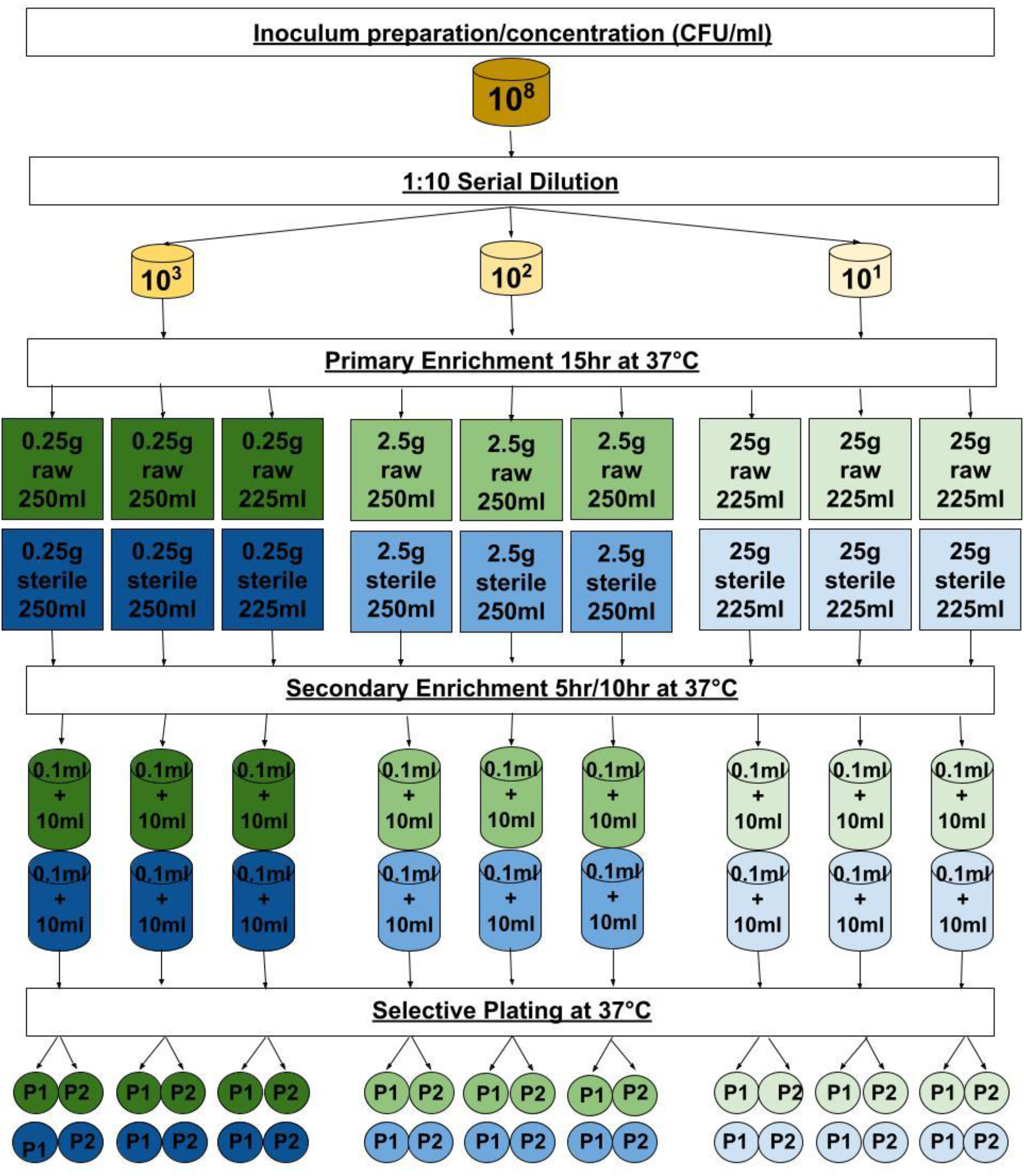
A workflow diagram showing the experimental design for simultaneous detection of multiple bacterial pathogens in soil. Each cylinder represents four tubes, one per selective enrichment broth; Each circle represents four plates (P1 & P2), one per selective growth; The green and blue color represents non-sterile and sterile soil samples, respectively, while the intensity of the color indicates the concentration of the spiked strains where the darkest color intensity represents the highest concentration (10^3); NS - non-sterile soil.

Following primary incubation, 100 µl of liquid culture from each of the bags was transferred into two falcon tubes containing 10 mL of respective selective enrichment broth for each pathogen. Rappaport-Vassiliadis Soya (RVS) broth (HiMedia India) was used to selectively enrich the growth of *S. flexneri* and *S. enterica*. Fraser broth (OXOID) and EC broth (OXOID) were chosen for the enrichment of *L. monocytogenes* and STEC, respectively. All the broths were prepared according to manufacturer’s instructions. The tubes containing selectively enriched aliquots were sealed, agitated and half of the tubes were incubated for 5 hr, and the other half were incubated for 10 hr. Fraser and E. coli broth cultures were incubated at 37°C, while the tubes containing RVS broth were incubated at 41.5°C. After the selective enrichment period, approximately 10 µl of enrichment was streaked onto two selective plates (XLD, MacConkey, PALCAM) corresponding according to pathogen type using a sterile inoculating loop. For reasons described in results, we shifted from use of PALCAM agar from HiMedia to PALCAM agar from OXOID in the rest of the experimental trials. All the plates were incubated for 24 hr at 37°C except for XLD which was incubated at 41.5°C.

Plates with growth of typical colony for the target pathogen(s) were recorded as positive. One morphologically distinct and typical colony from each matrix of soil and bacterial concentration were subcultured into Tryptic Soya Broth (TSB, OXOID, UK) and incubated at 37°C for 24 hr for preservation at -20°C in 0.5 ml TSB and 1 ml of 80% glycerol until used for molecular confirmation of pathogen presence.

### Molecular Detection of Pathogens

The presumptive pathogen isolates from the multi-pathogen soil spiking experiment were tested by quantitative real time PCR (qRT-PCR) using pathogen-specific assays to confirm colonies matching the expected phenotype on plates with the spiked known pathogens. Briefly, one sterile loopful from previously preserved colonies was added to 10 mL of LB nutrient broth in 15 mL Falcon tubes and incubated at 37°C for 24 hr. After incubation, 1 mL of the liquid culture from each tube was centrifuged for 2 minutes at 3000 RPM, the supernatant discarded, and the bacterial pellet was re-suspended in 100 µl of DNA/RNA free water. The tubes were then boiled at 100°C for 10 minutes, centrifuged at high speed (15,000 rpm) for 1 minute, transferred the supernatant containing bacterial DNA into new sterile tubes and preserved at – 20°C until used for qRT-PCR analysis. If growth was observed on process negative controls, colonies were also selected for pathogen testing.

The RT-qPCR assay was performed using TaqMan fast advanced Master Mix (Applied Biosystems, USA), Taqman Assay containing target primers and probes, cDNA template and nuclease free water (Inc, 2021). The primer and probe sequences used for the RT-qPCR are referenced elsewhere.^36^ The PCR was performed in a QuantStudio 7 Pro with run conditions of incubation at 50°C for 2 minutes, denaturing at 95°C for 20 seconds, and 40 cycles of 95°C for 1 second, and annealing/extension at 60°C for 20 seconds.^37^ PCR negative controls were performed using nuclease free water in place of sample DNA. Positive controls were obtained by growing the previously spiked known strains on selective media followed by DNA extraction using the same boiling method. Primary detection results are described in bacterial CFU per gram of soil for comparison.

### Process Evaluation of Protocol in Naturally Sourced Soils

A process evaluation was conducted at Maseno University in Kisumu, Kenya to assess the feasibility of the protocol in normal workflow laboratory settings as well as its effectiveness at each step of the processing pipeline at recovering one or more target bacterial species in environmental samples collected in a community with high pathogen transmission rates. Environmental samples collected and processed for the Pathogen Transmission and Health Outcome Models of Enteric Disease (PATHOME) study were used for field testing the protocol.^36^ The PATHOME study is a longitudinal study in Kenya that collected longitudinal data of children, their caregivers and their environment through a 14-day period with an aim of estimating the presence and concentrations of these common enteric pathogens from infants’ and animals’ feces, caregiver’s hands, infant’s toys, household and community soil, household drinking water, and community’s surface water samples. For field testing of the above methodology, 23 soil samples (7 household, 16 public domain) were selected from the PATHOME biorepository. Soil samples were a 20-gram composite of soil from different locations in the household or a swab of a 25 cm squared floor space. The protocol tested was ultimately modified to accommodate laboratory workflow limitations, specifically the incubation times for primary enrichment and secondary enrichment were increased to 24 hours.^36^

To evaluate how the protocol influenced microbial recovery in each sample, an aliquot of each sample was retrieved from four important sample processing steps: pre-enrichment (100 uL), primary enrichment (100 uL), selective secondary enrichment (100 uL), and selective plating (pool of up to 10 CFU resuspended in 100 uL BPW). DNA was extracted using a DNA/RNA mini-pathogen prep kit (Fisher Scientific, Waltham, MA) and tested by qRT-PCR for *S. enterica*, Shigella spp., *L. monocytogenes*, and STEC *E. coli* using the protocol described above. In addition, DNA was tested for genes of Enteropathogenic (EPEC), Enteroaggregative (EAEC), and Enterotoxigenic (ETEC) since these types of *E. coli* have previously been detected in soil from these communities,^12^ and could be enriched and isolated by the same protocol steps. DNA was further tested for *Campylobacter jejuni* to explore whether these bacteria could be detected. The protocol for soil samples was also tested on 15 water samples, 7 hand rinse samples, 7 toy rinse samples from the PATHOME biorepository to evaluate protocol usefulness for other sample types.

### Data Management and Analysis

The cycle thresholds (CTs) of qPCR results for each gene assay were set at 35 CT to classify an amplification result as “positive” or “negative”, with two (ETEC_LT set at 29.05, and ETEC_Sth set at 32) assays set at lower thresholds. The thresholds were determined by running negative controls, prior to, and alongside the samples, and then using a cut-off that was 95% of the lowest false-positive value. A false positive in a PCR negative control required retesting. Duplicate sets of tests for each pathogen gene and sample were evaluated for consistency. For *S. enterica (ttr), L. monocytogenes (hipA)*, and *C. jejuni/coli (cadF)*, a single gene assay was used for detection and determination of soil positivity. For *Shigella* spp. (*ipaH* or *virG)*, EPEC (*bfpA* or *ae*), ETEC (*elt* or *est*), STEC (*stx1* or *stx2*), or EAEC (*ataA* or *aaiC*) a positive result for either gene indicator was considered a positive result for the type of pathogenic bacteria. Positive detections are reported as counts per total samples tested.

Conclusions about detection rates across steps of sample processing are based upon observable increases or decreases in counts by species and across species.

## RESULTS

### Confirmation of single pathogen recovery from spiked samples

Pathogens were recovered from all sterilized 2.5 g soil samples spiked with the highest 10^3^ CFU concentration of each bacteria strain after 3 and 5 hours of primary enrichment (Table 1). When pre-enriched for 5 hours, all pathogens were consistently recovered from all four spiked replicate samples at concentrations as low as 10^1^ CFU/2.5 g bag, while the recovery rate for *S. flexneri* and *L. monocytogenes* was 3 of 4 replicates at the lowest concentration. The detection of *S. enterica* and *S. flexneri* became inconsistent at 10^2^ CFU spiking concentration when incubated for a 3-hour period. In the first few trials, *L. monocytogenes was observed to* be frequently overgrown by unidentified microorganisms with indistinct morphology on PALCAM agar obtained from HiMedia. Further trials comparing the growth of *L. monocytogenes* between PALCAM agar from HiMedia versus Oxoid manufacturers found typical colony morphology of *L. monocytogenes* without background growth on the negative control plates prepared from the OXOID PALCAM agar. This was observed in every trial except in the case of the growth on the negative control plates for the sterile soil samples, in which the growth of unknown microorganisms was observed on both plates prepared from the agars. Nonetheless, the experiment demonstrated that the growth was not attributed to environmental factors or experimental errors. These experiments confirmed that all pathogens could be recovered from low initial spiking concentrations using just 5 hr of primary enrichment incubation but suggested longer incubation times may be needed for reproducible detection of *S. flexneri* and *L. monocytogenes*.

**Table 1.**
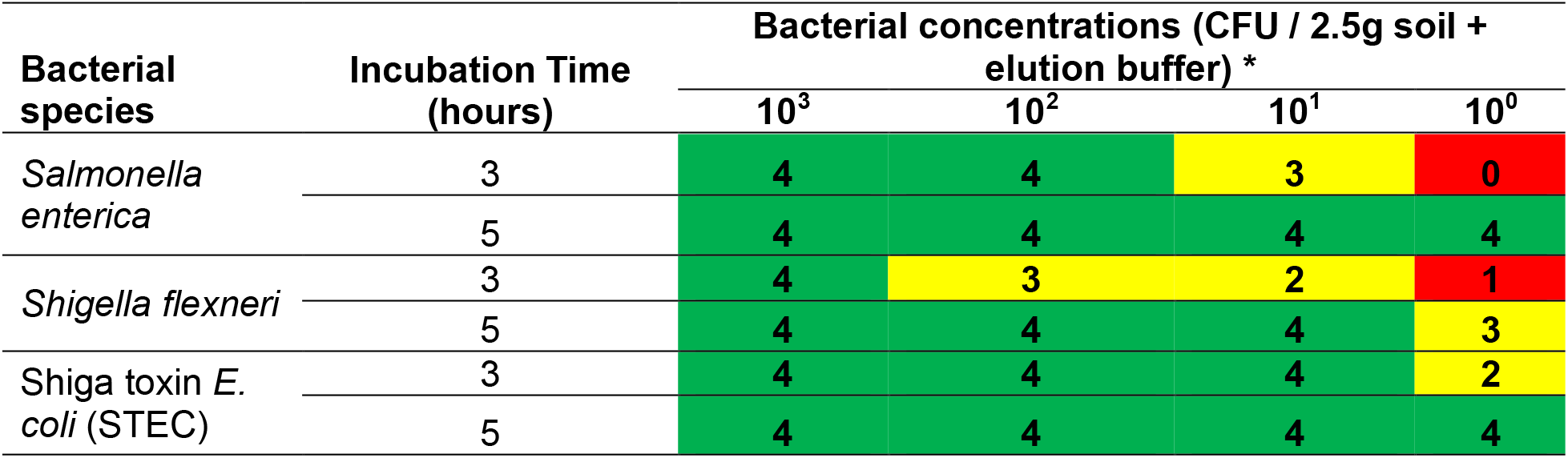
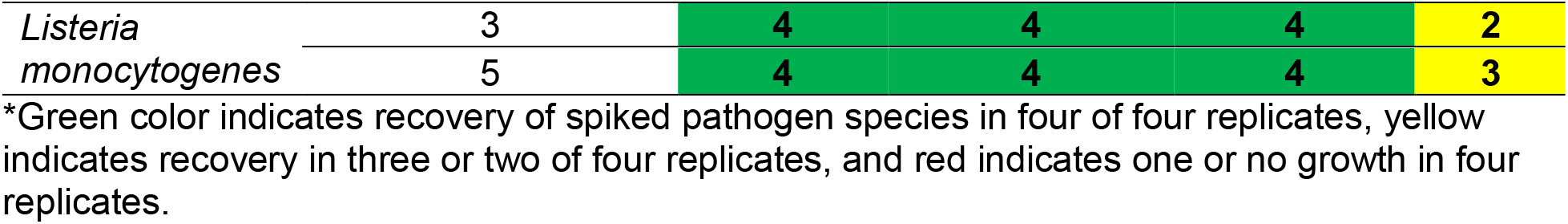
Frequency of recovery of single pathogen from sterile soil without secondary selective enrichment step at two different primary enrichment incubation periods.

### Consistency in Pathogen Co-detection after Secondary Enrichment

In the multi-pathogen spiking experiments, primary enrichment time was increased to 15 hr to promote stronger growth of *S. flexneri* and *L. monocytogenes* and to add a secondary selective enrichment step to determine if non-specific growth of soil microbiota could be reduced or eliminated.^38-42^ All pathogen types were recovered from soil spiked with pooled cultures of *S. enterica, S. flexneri*, STEC and *L. monocytogenes* bacteria at varying concentrations per gram of soil ratios and secondary enrichment conditions. In all trials, qPCR confirmed a 100% recovery rate for STEC and *L. monocytogenes* in both sterile (Table 2A) and non-sterile soil samples (Table 2B) irrespective of the sample quantity, pathogen concentrations and secondary enrichment incubation times. *S. enterica* was recovered in most replicates in both soil types, with consistent recovery observed in sterile soil with a bacterial concentration 10^1^ CFU/sample (equivalent to 10^-1^/g to 10^1^ per volume). The reduction in reproducible recovery below 10^1^ CFU across soil volumes rather than CFU per gram indicated that protocol performance was influenced most by how much pathogen was sampled, regardless of sample volume. A similar decline in reproducible replicate recovery per CFU spiked was observed for *S. enterica* in non-sterile soil albeit at a tenfold higher bacterial concentration (10^2^ CFU/sample), with CFU/g ratio having little influence on protocol efficacy. The 10 hr secondary enrichment slightly increased detection rates at lower concentration per gram ratios over 5 hr secondary enrichment. Contrary to the results of the single pathogen recovery, the detection rate of *S. flexneri* was about 50% on average and was not influenced by soil sample volume, pathogen spiking concentration or enrichment time. Surprisingly, the reproducibility of *S. flexneri* recovery was better, albeit marginally, in non-sterilized soil although detection was not improved with increases in CFU per soil/buffer mixture or CFU/g. The 15 hr primary enrichment, 10 hr secondary enrichment and PCR confirmation of isolated colonies was determined to be an acceptable minimum set of conditions for co-recovery of STEC, *L. monocytogenes*, and *S. enterica* of 10^1^ CFU overall concentration or as low as 1 CFU per gram, although potentially suboptimal for *S. flexneri* relative to just primary enrichment.

**Table 2.** Frequency of recovery of multiple enteric bacteria from sterile (A) and non-sterile (B) soil using secondary selective enrichment culture methods at two different incubation periods and varied bacteria to soil volume concentrations.

| A. SPIKED STERILE SOIL |  |  |  |  |  |  |  |  |  |  |
| --- | --- | --- | --- | --- | --- | --- | --- | --- | --- | --- |
| Bacteria | Incubation Time<br>(hours) | Pathogen CFU per soil volume |  |  |  |  |  |  |  |  |
|  |  | 0.25 g |  |  | 2.5 g |  |  | 25 g |  |  |
|  |  | 10<sup>2</sup> | 10<sup>1</sup> | 10<sup>0</sup> | 10<sup>2</sup> | 10<sup>1</sup> | 10<sup>0</sup> | 10<sup>2</sup> | 10<sup>1</sup> | 10<sup>0</sup> |
| *Salmonella enterica* <sup>1</sup> | 5 | 4 | 4 | 3 | 4 | 4 | 2 | 4 | 4 | 2 |

|  | 10 | 4 | 4 | 4 | 4 | 4 | 2 | 4 | 4 | 2 |
| --- | --- | --- | --- | --- | --- | --- | --- | --- | --- | --- |
| *Shigella flexneri*<sup>2</sup> | 5 | 1 | 1 | 0 | 0 | 1 | 0 | 1 | 1 | 0 |
|  | 10 | 0 | 0 | 0 | 1 | 1 | 1 | 4 | 0 | 0 |
| Shiga toxin *E. coli* (STEC)<sup>3</sup> | 5 | 4 | 4 | 4 | 4 | 4 | 4 | 4 | 4 | 4 |
|  | 10 | 4 | 4 | 4 | 4 | 4 | 4 | 4 | 4 | 4 |
| *Listeria monocytogenes*<sup>4</sup> | 5 | 4 | 4 | 4 | 4 | 4 | 4 | 4 | 4 | 4 |
|  | 10 | 4 | 4 | 4 | 4 | 4 | 4 | 4 | 4 | 4 |
| **B. SPIKED NON-STERILE SOIL** |  |  |  |  |  |  |  |  |  |  |
| Bacteria | Incubation Time (hours) | Pathogen CFU per soil volume |  |  |  |  |  |  |  |  |
|  |  | 0.25g |  |  | 2.5g |  |  | 25g |  |  |
|  |  | 10<sup>2</sup> | 10<sup>1</sup> | 10<sup>0</sup> | 10<sup>2</sup> | 10<sup>1</sup> | 10<sup>0</sup> | 10<sup>2</sup> | 10<sup>1</sup> | 10<sup>0</sup> |
| *Salmonella enterica*<sup>1</sup> | 5 | 4 | 3 | 2 | 4 | 2 | 2 | 4 | 2 | 2 |
|  | 10 | 4 | 4 | 2 | 4 | 2 | 2 | 4 | 3 | 2 |
| *Shigella flexneri*<sup>2</sup> | 5 | 0 | 2 | 2 | 3 | 2 | 2 | 3 | 4 | 2 |
|  | 10 | 0 | 2 | 4 | 2 | 4 | 1 | 2 | 3 | 2 |
| Shiga toxin *E. coli* (STEC)<sup>3</sup> | 5 | 4 | 4 | 4 | 4 | 4 | 4 | 4 | 4 | 4 |
|  | 10 | 4 | 4 | 4 | 4 | 4 | 4 | 4 | 4 | 4 |
| *Listeria monocytogenes*<sup>4</sup> | 5 | 4 | 4 | 4 | 4 | 4 | 4 | 4 | 4 | 4 |
|  | 10 | 4 | 4 | 4 | 4 | 4 | 4 | 4 | 4 | 4 |

\*Green color indicates recovery of spiked pathogen species in four of four replicates, yellow indicates recovery in three or two of four replicates, and red indicates one or no growth in four replicates.
<sup>1</sup> *S. enterica* spiking concentrations 4.8 x 10<sup>2</sup>, 4.8 x 10<sup>1</sup>, 4.8 x 10<sup>0</sup>.
<sup>2</sup> *S. flexneri* spiking concentrations 5.3 x 10<sup>2</sup>, 5.3 x 10<sup>1</sup>, 5.3 x 10<sup>0</sup>.
<sup>3</sup> STEC spiking concentrations 5.2 x 10<sup>2</sup>, 5.2 x 10<sup>1</sup>, 5.2 x 10<sup>0</sup>.
<sup>4</sup> *L. monocytogenes* spiking concentrations 5.5 x 10<sup>2</sup>, 5.5 x 10<sup>1</sup>, 5.5 x 10<sup>0</sup>.

### Process Evaluation of a Protocol in a Field Study of Soil from Neighborhoods with Prior Soil Contamination

Primary and secondary enrichment times had to be expanded to 24 hr to be compatible with sample collection to processing workflow, but pathogen detection overall in soil collected in Kenya was high and did not appear to be impeded by the longer enrichment time. The process evaluation of the protocol found that EPEC were detected at high rates at the onset of processing, with STEC, ETEC, and *Shigella spp*. detected at lower rates, and no *S. enterica, L. monocytogenes*, or *C. jejuni/coli* (Table 3). Primary enrichment for 24 hr in BPW enhanced EPEC and STEC qPCR detection rates and led to detection of 2 *S. enterica* positive samples. The chances of detection for all *E. coli* and *S. enterica* were highest when qPCR testing DNA extracted after selective secondary enrichment, before selective plating. In contrast to the controlled spiking experiments, testing pools of 5 to 10 colonies isolated from plated secondary enrichment greatly reduced pathogen detection. Conducting qPCR testing of DNA extracted from secondary enrichment was the optimal protocol, shifting positivity rates from onset to detection by 50% to 91% (EPEC), 0% to 43% (EAEC), 17% to 65% (STEC), 4% to 65% (ETEC), and 0% to 52% (*S. enterica*). *Shigella spp*. were detected at onset and primary enrichment but not after secondary enrichment, so evidence suggests a multi-pathogen soil testing protocol should include a step for DNA extraction of primary enrichment for *Shigella spp*. testing prior to secondary enrichment.

**Table 3.** Quantitative PCR detection of enteric pathogens in soil samples at key steps in the culture process (N = 23)

|  | Enrichment onset | Primary enrichment | Secondary enrichment | Selective plating |
| --- | --- | --- | --- | --- |
| EPEC | 12 | 18 | 21 | 2 |
| EAEC | 0 | 0 | 10 | 0 |
| STEC | 4 | 12 | 15 | 3 |
| ETEC | 1 | 1 | 15 | 2 |
| Salmonella | 0 | 2 | 12 | 6 |
| Shigella | 1 | 1 | 0 | 0 |

\* *L. monocytogenes* and *C. jejuni* were not detected among the selected soil samples.

## DISCUSSION

Exposure to contaminated soil is an important transmission pathway for bacterial enteric diseases, especially in children.^5,8,11,15,16^ Co-occurrence of bacterial enteric pathogens in soil in settings impacted by fecal waste makes microbiological testing for multiple common bacteria pathogens in soil important for health risk assessment of the natural environment.^11,12^ This study demonstrated that a streamlined primary and secondary enrichment process with PCR validation can consistently and simultaneously detect low concentrations of *S. enterica, L. monocytogenes*, and multiple types of pathogenic *E. coli* in spiked (Iowa) or naturally contaminated (Kenya) soil. While we detected low concentrations per gram of *Shigella* spp., replicates were critical to ensure detection and the optimal timing for detection was best after primary enrichment, rather than selective secondary enrichment. By reducing reliance on selective plating and allowing several pathogens to be assessed from shared enrichment steps, the protocol may reduce consumable use, hands-on time and overall turnaround time. Formal cost and workflow evaluations are still required. Our study also aimed to shorten the time required for pathogen detection. Initial experimental assays in Iowa suggested that a 15-hour primary and 5-hour secondary enrichment followed by 24-hour selective agar culture and PCR testing of colonies (∼48 hours) would be sensitive for most bacteria. While this protocol shifted, the final protocol involving 24-hour primary and 24-hour secondary enrichment with PCR testing of secondary enrichment (52 hr) remained one or more days shorter than many protocols involving selective plating. For *Shigella* spp. the testing at the primary enrichment stage shortens detection timelines by at least two days.

Our process began with controlled experiments that verified the minimum set of conditions for detecting each pathogen species in microbially sterile soil, then non-sterile soils containing native microbes that would be expected in real-world testing conditions. Incubating environmental samples for six or more hours in universal pre-enrichment broth can improve the recovery of bacterial pathogens.^43^ PCR testing after 5 hours of primary enrichment was faster and more efficient for *S. enterica* detection in food compared to the longer incubation methods, but has not been tested in soil nor with pathogen species mixtures.^35^ Our spiking experiments showed reproducible 100% detection of 10^1^ CFU or lower concentrations for *S. enterica, L. monocytogenes, S. flexneri* and STEC individually spiked in sterilized soil using just 5 hrs primary enrichment and plating. This agrees with other studies that demonstrated early detection of Salmonella,^44-47^ STEC,^48^ L. monocytogenes,^47,49^ and Shigella^50^ in different food samples spiked with known strains after 3 to 6 hr, 3 hr, 5 hr, and 6 hr enrichment periods, respectively.

Prior studies have described using short non-selective primary enrichment of food samples to detect multiple pathogens. For instance, *Salmonella* spp., *E. coli* O157:H7, and *L. monocytogenes* were co-detected in dairy products pre-enriched in Tryptic Soy Broth for 7 hours without selective enrichment.^47,49^ *Salmonella* spp. and *E. coli* O157:H7 were also detected in ground beef and chicken breast pre-enriched in BPW for 3 hours.^48^ Soil poses unique challenges with pathogen species recovery and differentiation compared to drinking water and food due to the presence of naturally occurring soil microbes that could interact with pathogens. Native microbes could outcompete certain types of slow growing bacteria, like *S. flexneri*.^51-54^ Other enteric bacteria like STEC can also inhibit *Shigella* growth.^55,56^ While a previous study indicated that selective broth enrichment did not improve the detection of diarrheagenic *E. coli*,^57^ a secondary enrichment step was added to determine if selective conditions could reduce non-specific growth of soil microbiota.^38-42^ Primary enrichment time was also increased to 15 hr to foster *S. flexneri (*and *L. monocytogenes)* growth. In contrast with other studies, the selective enrichment step lowered the detection limit for STEC, as well as *L. monocytogenes*, by one log to 10^0^ CFU (any soil volume) but increased detection limits by a log for *S. enterica. S. flexneri* detection rates diminished when mixed with other pathogens and were not improved with additional growth on secondary enrichment media. Recovery of *Shigella* from the environment may be a sustained challenge if it co-occurs as a contaminant alongside other pathogens. A 15 hr primary enrichment of soil followed by a pathogen-specific selective enrichment step of 10 hours and PCR validation was optimal for multi-pathogen recovery of concentrations as low as 1 to 10 CFU in soil.

The protocol was field tested in a community where detection of these pathogens in soil is common, and a process evaluation was used to investigate how each step of the protocol affected detection rates for our pathogen targets.^12^ The process evaluation confirmed what the experimental work had shown: (1) that primary enrichment improves growth and detection of all of our target bacteria; (2) that a single primary enrichment can be used for co-detection of multiple *Enterobacteriaceae* pathogens; and (3) under the conditions evaluated, selective secondary enrichment did not improve Shigella detection and may have reduced sensitivity relative to testing after primary enrichment. It also revealed some surprises, such as the very high loss in sensitivity when microbial validation tests were performed by testing colonies from selective agar. Testing secondary enrichment media, or primary enrichment for *Shigella spp*. led to significant increases in detection rates, reduced pathogen detection time by at least one day, and reduced the costs associated with preparing and using the additional consumable supplies. Furthermore, processing of surface water with the same protocol generated consistent results and suggested that this protocol could be applied to a variety of sample types.

This study had some limitations. First, the methods were evaluated using only two soil types from Iowa and Kenya; therefore, their performance may differ in settings with distinct physical, chemical and microbiological soil characteristics. Second, the low prevalence and inconsistent recovery of *Shigella* spp. across experimental replicates limited our ability to determine the optimal enrichment and detection conditions for this pathogen. Methodological changes introduced during the study, including the replacement of culture media for *L. monocytogenes*, may also have affected comparability across experiments. In addition, autoclave-sterilized soil may not have provided a completely sterile or representative matrix because some microorganisms may have survived sterilization or been introduced during subsequent handling. Enrichment times were also adjusted to accommodate field-sample collection and laboratory-processing schedules, which may have influenced pathogen recovery and turnaround time.

Future protocol optimization could explore ways to shorten enrichment times to reduce total processing time as well as investigate optimal testing points for the detection of *S. flexneri* in soil. As noted, we changed *L. monocytogenes* media during the study due to observation of visible growth of morphologically indistinct organisms in non-spiked sterile soil samples for the negative control on OXOID PALCAM agar. Breakthrough growth on PALCAM could have been due to varying efficacy of autoclaves as a method for sterilizing natural soil bacteria, post-sterilization contamination, breakthrough growth of non-target organisms, or limited sensitivity or colony-morphology specificity of PALCAM agar.^58,59^

## CONCLUSIONS

Overall, the findings support a shared enrichment and PCR-based approach for detecting several enteric bacterial pathogens in soil. The method performed consistently for *S. enterica, L. monocytogenes* and pathogenic *E. coli*, while *Shigella* detection remained less reliable and was most effective after primary enrichment. Further validation across diverse soil types, larger numbers of naturally contaminated samples and additional laboratory settings, and protocol timepoints will reveal whether this method can improve enteric pathogen exposure assessment for soil.

## Data Availability

All data produced in the present work are contained in the manuscript with raw data available upon reasonable request to the authors

## AUTHOR CONTRIBUTIONS

Conception and funding acquisition: K.K.B., D.S.; design: A.K., M.S., F.D.G., C.O. and K.K.B.; sample collection and laboratory analysis: A.K., M.S., B.O., J.A., C.S.A., F.D.G.; visualization: A.K., E.M.M; data curation: E.M.M; original draft preparation: A.K., M.S., F.D.G., E.M., K.K.B.; manuscript review and editing, K.K.B, E.M.M.; supervision and project administration, K.K.B., C.O., S.S. All authors have read and agreed to the published version of this manuscript.

## DATA AVAILABILITY STATEMENT

The authors confirm that the necessary information to replicate the outcomes of this study is available in the manuscript.

