## Supplemental File 1 for "Design and evaluation of a streamlined enrichment method for faster co-detection of enteric bacterial pathogens in natural soils and environmental samples"

Supplemental Table 1: Pathogen detection by monoplex qPCR assays in surface water samples at key steps in culture process (N = 8)

| | Enrichment onset | Primary enrichment | Secondary enrichment | Selective plating |
|---|---|---|---|---|
| EPEC | 2 | 7 | 8 | 0 |
| EAEC | 0 | 0 | 6 | 0 |
| STEC | 3 | 8 | 8 | 0 |
| ETEC | 0 | 0 | 1 | 1 |
| *Salmonella enterica* | 0 | 2 | 4 | 1 |
| *Shigella* spp. | 0 | 3 | 0 | 0 |
| *Campylobacter jejuni/coli* | 0 | 0 | 0 | 0 |
| *Listeria monocytogenes* | 0 | 0 | 0 | 0 |

- Enteropathogenic *Escherichia coli* (EPEC); Enteroaggregative *Escherichia coli* (EAEC); Shiga toxin-expressing *Escherichia coli* (STEC); Enterotoxigenic *Escherichia coli* (ETEC). Gene targets provided in supplement.

Supplemental Table 2: Pathogen detection by monoplex qPCR assays in drinking water samples at key steps in culture process (N = 7)

| | Enrichment onset | Primary enrichment | Secondary enrichment | Selective plating |
|---|---|---|---|---|
| EPEC | 0 | 0 | 0 | 0 |
| EAEC | 0 | 0 | 0 | 0 |
| STEC | 0 | 0 | 0 | 0 |
| ETEC | 0 | 0 | 0 | 0 |
| *Salmonella enterica* | 0 | 0 | 0 | 0 |
| *Shigella* spp. | 2 | 3 | 0 | 0 |
| *Campylobacter jejuni/coli* | 1 | 0 | 0 | 0 |
| *Listeria monocytogenes* | 0 | 0 | 0 | 0 |

- Enteropathogenic *Escherichia coli* (EPEC); Enteroaggregative *Escherichia coli* (EAEC); Shiga toxin-expressing *Escherichia coli* (STEC); Enterotoxigenic *Escherichia coli* (ETEC). Gene targets provided in supplement.

Supplemental Table 3: Pathogen detection by monoplex qPCR assays in hand-rinse samples at key steps in culture process (N = 7)

| | Enrichment onset | Primary enrichment | Secondary enrichment | Selective plating |
|---|---|---|---|---|
| EPEC | 0 | 0 | 0 | 0 |
| EAEC | 0 | 0 | 0 | 0 |
| STEC | 0 | 0 | 0 | 0 |
| ETEC | 0 | 0 | 0 | 0 |
| *Salmonella enterica* | 0 | 0 | 0 | 0 |
| *Shigella* spp. | 1 | 0 | 0 | 0 |
| *Campylobacter jejuni/coli* | 0 | 0 | 0 | 0 |
| *Listeria monocytogenes* | 0 | 0 | 0 | 0 |

- Enteropathogenic *Escherichia coli* (EPEC); Enteroaggregative *Escherichia coli* (EAEC); Shiga toxin-expressing *Escherichia coli* (STEC); Enterotoxigenic *Escherichia coli* (ETEC). Gene targets provided in supplement.

Supplemental Table 4: Pathogen detection by monoplex qPCR assays in toy-rinse samples at key steps in culture process (N = 7)

| | Enrichment onset | Primary enrichment | Secondary enrichment | Selective plating |
|---|---|---|---|---|
| EPEC | 0 | 2 | 2 | 0 |
| EAEC | 0 | 0 | 0 | 0 |
| STEC | 0 | 0 | 0 | 0 |
| ETEC | 0 | 2 | 1 | 0 |
| *Salmonella enterica* | 0 | 0 | 0 | 0 |
| *Shigella* spp. | 0 | 0 | 2 | 0 |
| *Campylobacter jejuni/coli* | 0 | 0 | 0 | 0 |
| *Listeria monocytogenes* | 0 | 0 | 0 | 0 |

- Enteropathogenic *Escherichia coli* (EPEC); Enteroaggregative *Escherichia coli* (EAEC); Shiga toxin-expressing *Escherichia coli* (STEC); Enterotoxigenic *Escherichia coli* (ETEC). Gene targets provided in supplement.
